# Deploying wearable sensors for pandemic mitigation

**DOI:** 10.1101/2022.02.07.22270634

**Authors:** Nathan Duarte, Rahul K. Arora, Graham Bennett, Meng Wang, Michael P. Snyder, Jeremy R. Cooperstock, Caroline E. Wagner

## Abstract

Wearable sensors can continuously and passively detect potential respiratory infections, before or absent symptoms. However, the population-level impact of deploying these devices during pandemics is unclear. We built a compartmental model of Canada’s second COVID-19 wave and simulated wearable sensor deployment scenarios, systematically varying detection algorithm accuracy, uptake, and adherence. With current detection algorithms and 4% uptake, we found that deploying wearable sensors could have averted 9% of second wave SARS-CoV-2 infections, though 29% of this reduction is attributed to incorrectly quarantining uninfected device users. Improving detection specificity and offering confirmatory rapid tests each minimized incorrect quarantines and associated costs. With a sufficiently low false positive rate, increasing uptake and adherence became effective strategies for scaling averted infections. We concluded that wearable sensor deployment can meaningfully contribute to pandemic mitigation; in the case of COVID-19, technology improvements or supporting measures are required to reduce social and economic costs to acceptable levels.

## INTRODUCTION

Infectious disease outbreaks can have devastating health and economic consequences. Effective public health strategies are crucial for limiting transmission and minimizing these harms. One approach to controlling viral spread during pandemics – a “Find, Test, Trace, Isolate” (FTTI) strategy – relies on identifying and quarantining infectious individuals^1^. However, the COVID-19 pandemic has demonstrated that FTTI systems reliant on lab-based tests are often limited by missed hidden infection chains, which result from presymptomatic and asymptomatic transmission, and slow test result turnaround times^2,3^. Digital contact tracing and frequent use of rapid tests have the potential to fill these gaps, but both approaches have faced implementation barriers: inadequate participation levels, concerns around privacy and feasibility, and limited test availability^4–6^.

Wearable sensors have already been established as tools to detect deviations from users’ physiological baselines^7^. Recent findings suggest that wearable sensors may also be able to detect infections caused by respiratory pathogens, including SARS-CoV-2, before or absent symptoms^8–10^. Alavi *et al*, for example, developed an algorithm that analyzes patterns in smartwatch-captured overnight resting heart rate and provides real-time alerts of potential presymptomatic and asymptomatic SARS-CoV-2 infection^10^. If such algorithms were widely deployed, wearable sensors could be promising tools for outbreak mitigation, enabling FTTI systems to more rapidly identify (and subsequently isolate) infectious individuals, particularly those without symptoms. Wearable sensors would also offer the unique benefit of passive monitoring, which minimizes required user engagement, and could operate in privacy-preserving fashion because sensor data would not need to be shared with a centralized database. With these potential benefits in mind, several studies have focused on developing wearable sensor-based infectious disease detection algorithms; however, to the best of our knowledge, the real-world impact of deploying these devices for pandemic mitigation has yet to be explored.

In this study, we assessed how wearable sensors can help reduce the burden of infection during a pandemic with the overarching goal of guiding future research investment and policy. To do so, we used COVID-19 as an example and explored counterfactual scenarios in which these devices were deployed to combat Canada’s second wave. We built a compartmental epidemiological model in which wearable devices notify users of potential infection and prompt users to seek a confirmatory lab-based test, self-isolating while waiting for the result. We aimed to (1) assess the baseline impact of deploying currently available detection algorithms, (2) investigate how detection accuracy and behavioural parameters influence this impact, and (3) explore a complementary strategy in which rapid antigen tests are used to confirm wearable-informed notifications of potential infection.

## METHODS

### Counterfactual Scenarios

We simulated Canada’s second COVID-19 wave (September 1, 2020 to February 20, 2021). This time window allowed us to capture the dynamics of wearable sensor deployment during an acute phase of the pandemic, prior to broad vaccine availability, and at a time when the technology would have been ready and deployable.

First, we considered a baseline scenario in which wearable device users can download an application with currently available detection algorithms^10^. Then, we investigated the impact of technology and behavioural parameters: detection sensitivity and specificity; uptake, defined as the proportion of the population that has downloaded the application and uses their wearable device often enough; and adherence, defined as the proportion of users who comply with recommended next steps after a positive notification. Finally, we explored a complementary intervention where wearable device users with a positive notification are offered a confirmatory rapid antigen test before they are prompted to seek a lab-based test and self-isolate.

### Model description

We built a compartmental model based on a *Susceptible, Exposed, Infectious, Removed* (*SEIR*) framework (Figure 1). We split the *Infectious* state into three sub-states: *Presymptomatic, Asymptomatic*, and *Symptomatic*. All infected individuals enter the *Presymptomatic* infectious state after a latency period following exposure; some go on to develop symptoms (*Symptomatic*) while others do not (*Asymptomatic*).

**Figure 1:**
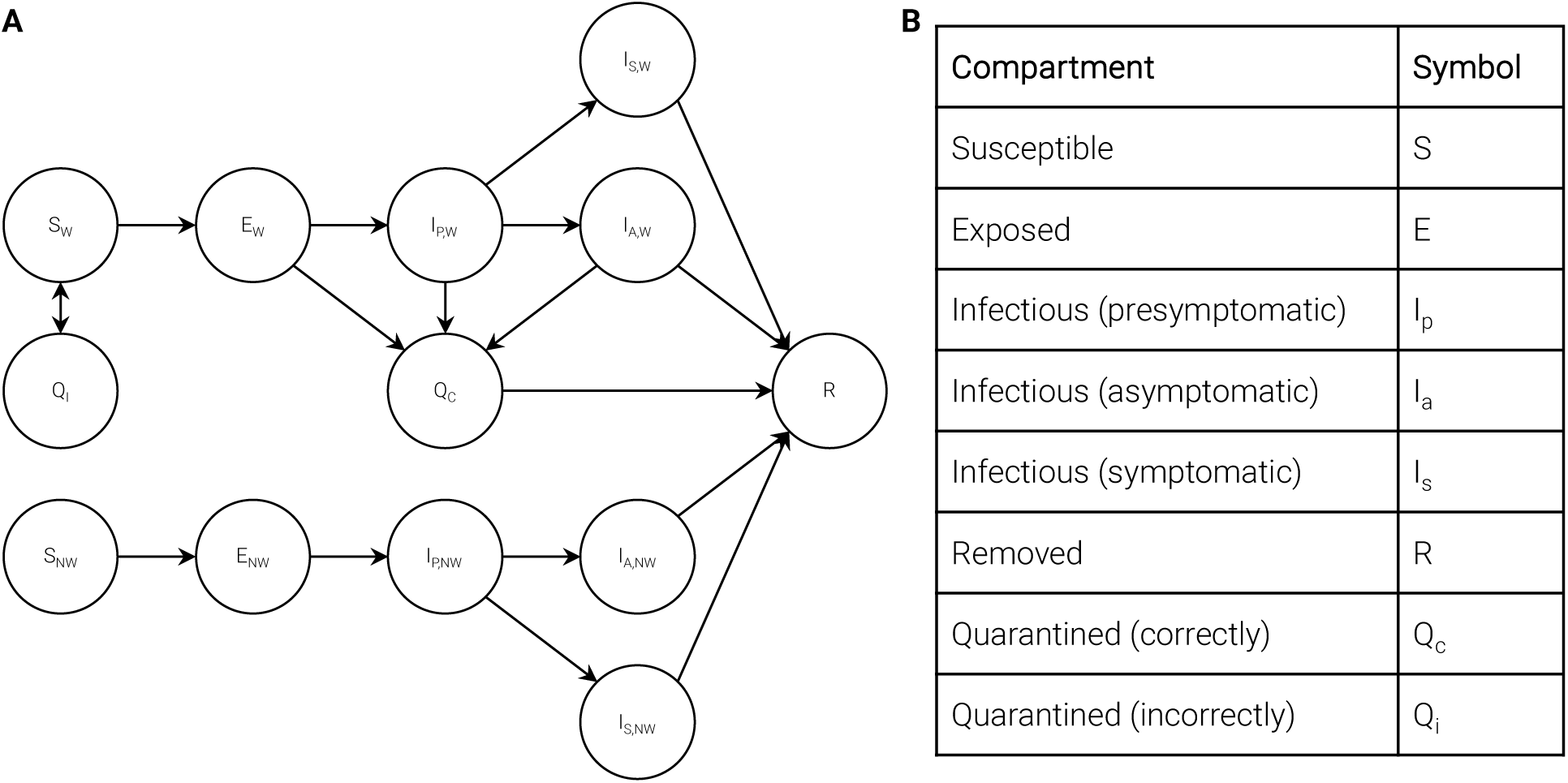
Compartmental model. Subscript “W” denotes a wearable user and “NW” indicates otherwise. Two key model equations are presented below; remaining parameters, equations, and assumptions are outlined in Supplementary Appendix Section 1.

To incorporate wearable sensor deployment, we stratified *Susceptible, Exposed*, and *Infectious* states by whether individuals are device users or not. Wearable device users can enter *Quarantined* states if they are notified of potential infection, and if they adhere to this notification by seeking a confirmatory lab-based test and self-isolating while awaiting the result. We made the simplifying assumption that users notified of potential infection either adhered to all recommended next steps or ignored the notification. We set the nominal lab-based test turnaround time to 2 days, and explored faster and slower turnaround times in Supplementary Appendix Section 5^11^. *Susceptible* users could be *Incorrectly Quarantined* due to a false positive notification and would re-enter the *Susceptible* state after receiving their lab-based test result. *Exposed* and *Infectious* device users would be *Correctly Quarantined* and would enter the *Removed* state (a longer quarantine until recovery) after their lab-based test confirms infection.

We assumed perfect lab-based test accuracy and did not account for potential reinfections, which were likely negligible during the simulation period^12^. We did not include a pathway for *Symptomatic* device users to enter *Quarantined* states because it is likely that a large fraction of these individuals already underwent some degree of self-isolation – behaviour accounted for in the historical transmission rate (β). In some scenarios, we also included a step where compliant users take a confirmatory rapid antigen test: if positive, we assumed they then take a lab-based test, self-isolating while awaiting the result; if negative, we assumed they return to normal behaviour. We investigated the impact of antigen test sensitivity in Supplementary Appendix Section 5.

### Simulation approach

To perform simulations, we first extracted the historical transmission rate (β) from the Institute for Health Metrics and Evaluation (IHME) infection model, a time series “nowcasting” model for incidence of infection in Canada (π)^13^. The IHME model estimates π by analyzing trends in confirmed cases, hospitalizations, and deaths; it validates the inferred π using seroprevalence data. We downloaded these data from the IHME website on December 7, 2021. We used modelled values for the incidence of infection because confirmed case counts are subject to incomplete case ascertainment and thereby underestimate the extent of infection^14^. We calculated β from π using Equation (1); this time series for the historical transmission rate incorporates all policy (e.g., restrictions, business closures, testing availability) and behavioural (e.g., adherence to restrictions, quarantines) measures that occurred. *N* represents the size of the entire population and λ represents the transmission potential in infected individuals without symptoms relative to those with symptoms.

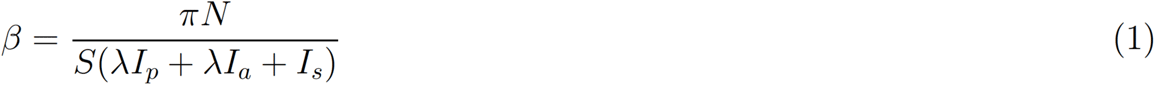

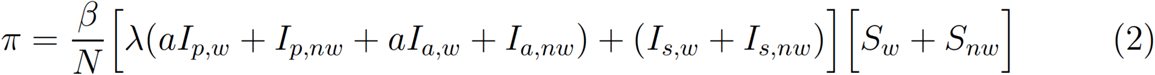

We then applied β according to Equation (2) to simulate counterfactual scenarios. Because some *Susceptible, Exposed*, and *Infectious* device users undergo self-isolation in the simulations, overall transmission decreases even though the transmission rate for all others remains unchanged. In Supplementary Appendix Section 5, we investigated the possibility that users not notified of potential infection act in a riskier fashion relative to historical behaviour^15^. *a* is a coefficient used to study this possibility and is nominally set to 1. Other model parameters, equations, and assumptions are presented in Supplementary Appendix Section 1.

We modeled asymptomatic prevalence (ρ), detection algorithm sensitivity (σ^w^) and specificity (ν^w^), and adherence (ψ) as beta-distributed random variables because these parameters were important sources of variance in our assessment of wearable sensors as pandemic mitigation tools. We sampled these variables and used the resulting values to generate an epidemic trajectory. We repeated this process 1,000 times, using these Monte Carlo simulations to model uncertainty in our estimates.

### Outcome measures

We calculated the number of averted infections and the percent reduction in the burden of infection to quantify the health impact of wearable sensor deployment. We measured the number of days incorrectly spent in quarantine per month per device user (a consequence of false positive notifications) as the primary indicator of the strategy’s social burden. To assess resource consumption, we quantified the number of additional lab-based tests (and rapid antigen tests, where applicable) required each day, on average. Finally, to provide more complete intuition around offering rapid antigen tests as a complementary intervention, we generated a first-order estimate of net healthcare expenditures in applicable scenarios, subtracting savings from costs. We approximated savings as the costs avoided by averting hospitalizations: we multiplied averted infections, the infection hospitalization ratio, and the average cost per hospitalization. We inferred the infection hospitalization ratio by dividing the total number of COVID-19 hospitalizations by the total number of infections during the simulation period. We approximated costs as expenditures required to fund lab-based tests and rapid antigen tests. Further details on this economic analysis are provided in Supplementary Appendix Section 3. We adhered to CHEERS guidelines for health economics cost benefit analyses^16^.

## RESULTS

### Baseline impact of wearable sensor deployment

We first investigated the baseline scenario in which detection algorithms that currently exist are made publicly available for device users to download and use (Figure 2)^10^. Upon notification of potential presymptomatic or asymptomatic infection, users are prompted to seek a confirmatory lab-based test, self-isolate while awaiting the result (nominally, for 2 days), and quarantine until recovery if positive. We used the nominal parameters in Supplementary Table 2, setting uptake, adherence, detection sensitivity, and detection specificity to 4%, 50%, 80%, and 92%, respectively.

**Figure 2:**
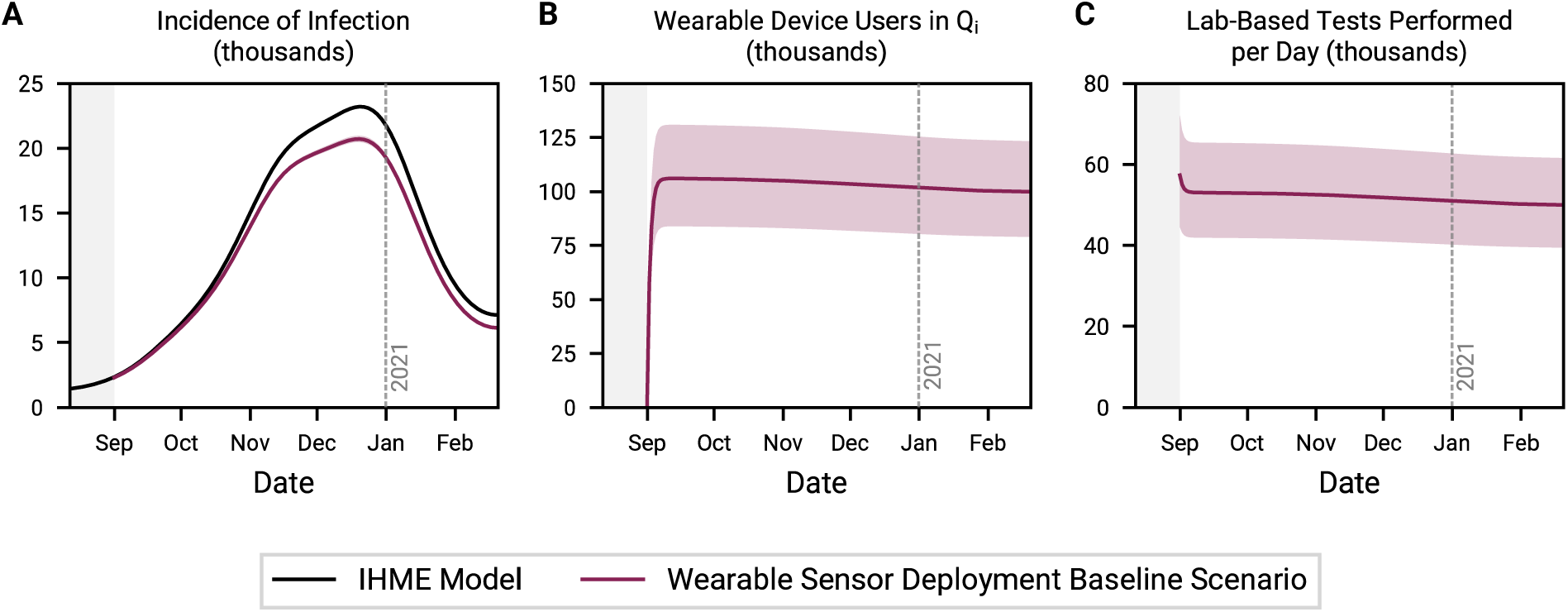
Baseline scenario for wearable sensor deployment. Time series depiction of the incidence of infection (A), the number of wearable device users incorrectly in quarantine (B), and the daily demand for lab-based tests (C). Uptake, adherence, detection sensitivity, and detection specificity are set to 4%, 50%, 80%, and 92%, respectively.

With 4% uptake, 219,700 (95% CI: 198,700–241,600) infections could have been averted during Canada’s second COVID-19 wave – a 9.4% (95% CI: 8.5–10.3%) reduction in the burden of infection. However, the social costs were high: between ∼75,000 and ∼125,000 device users were incorrectly self-isolating on any given day (Figure 2b). Moreover, between ∼40,000 and ∼65,000 additional lab-based tests were required each day (Figure 2c), corresponding to a 51.8% (95% CI: 41.0–64.0%) increase in demand relative to historical volumes. Historically, ∼101,000 lab-based tests were performed each day, on average, during the simulation time frame^13,17^. The number of individuals incorrectly in quarantine and daily demand for lab-based tests were steady over time because they depend on the number of *Susceptible* device users, adherence, and detection specificity; the gradual decrease can be attributed to the flow of users into the *Removed* state.

### Tradeoff between detection algorithm sensitivity and specificity

After their initial release on technology platforms, health detection algorithms can be updated and improved as more real-world data are collected. However, it is often challenging to dramatically raise detection sensitivity and specificity at the same time. We explored the implications of this tradeoff (Figure 3), varying detection sensitivity and specificity while keeping uptake and adherence constant at 4% and 50%, respectively.

**Figure 3:**
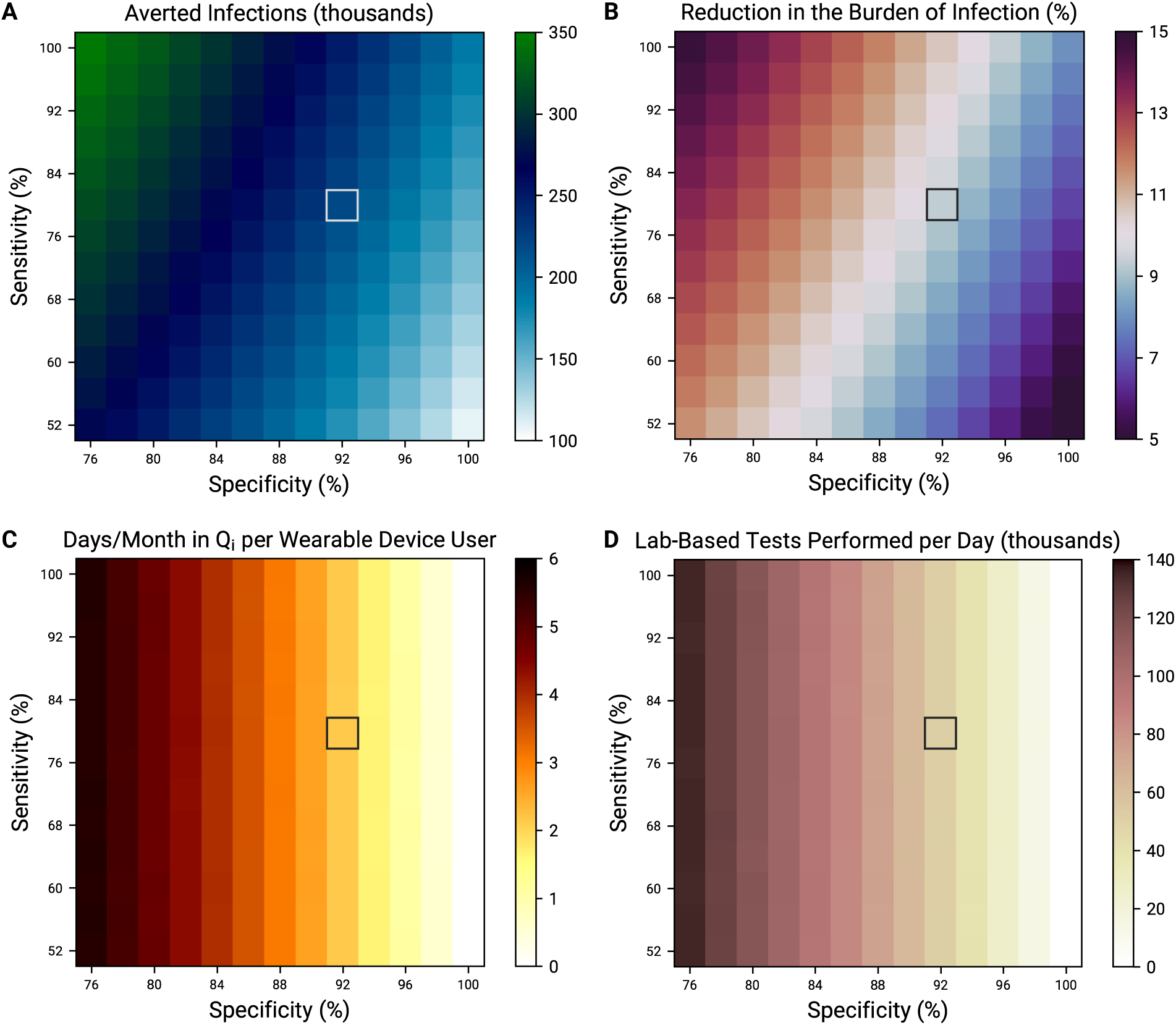
Tradeoff between detection sensitivity and specificity. Averted infections (A), reduction in the burden of infection (B), days incorrectly spent in quarantine per month per device user (C), and average daily demand for lab-based tests (D), all over the entire simulation period, as a function of detection sensitivity and specificity. Grey boxes denote nominal sensitivity (80%) and specificity (92%).

Increasing detection sensitivity increased the number of averted infections by prompting more *Infectious* users to self-isolate (Figures 3a and 3b). On the other hand, increasing specificity had a two-part effect. First, as specificity approached 100%, the number of days incorrectly spent self-isolating approached zero (Figure 3c); sensitivity had negligible impact on incorrect quarantines. Second, by virtue of decreasing the number of incorrect quarantines, increasing specificity resulted in a larger pool of *Susceptible* individuals; in turn, fewer infections were averted. However, despite this second effect, incorrect quarantines were not central to the strategy’s public health impact. In the baseline scenario presented above (80% detection sensitivity, 4% uptake, and 50% adherence), a 6.7% (5.9–7.3%) reduction in the burden of infection was still achievable with perfect detection specificity. As well, while 28.6% (95% CI: 18.3–38.2%) of averted infections could be attributed to incorrect quarantines in the baseline scenario, this proportion decreased as sensitivity improved (Supplementary Figure 4).

In theory, increasing detection sensitivity would increase demand for lab-based tests; however, this effect paled in comparison to the number of lab-based tests prompted by false positive notifications (Figure 3d). Lab-based test demand expectedly decreased as detection specificity increased.

### Impact of increasing uptake

Ensuring that public health measures reach sufficient levels of uptake has been a continued challenge through the COVID-19 pandemic. Digital contact tracing and vaccination efforts have demonstrated that well-constructed policies – for example, incentivizing participation – can improve uptake of measures^18,19^. Here, we explored the role of uptake to provide relevant context for the design of wearable sensor deployment policies (Figure 4; Supplementary Figure 1). We estimated that uptake would fall between 0.5% and 7.5% (Supplementary Tables 2-4) at baseline but chose to present outcomes at all levels of uptake to illustrate emergent phenomena. We also explored multiple technology scenarios, setting “high” detection sensitivity and specificity at 96.0% and 98.4%, respectively; we based these increases on the respective goals of capturing 20% more infections and reducing the false positive rate by 80% relative to nominal values. We kept adherence constant at 50%.

**Figure 4:**
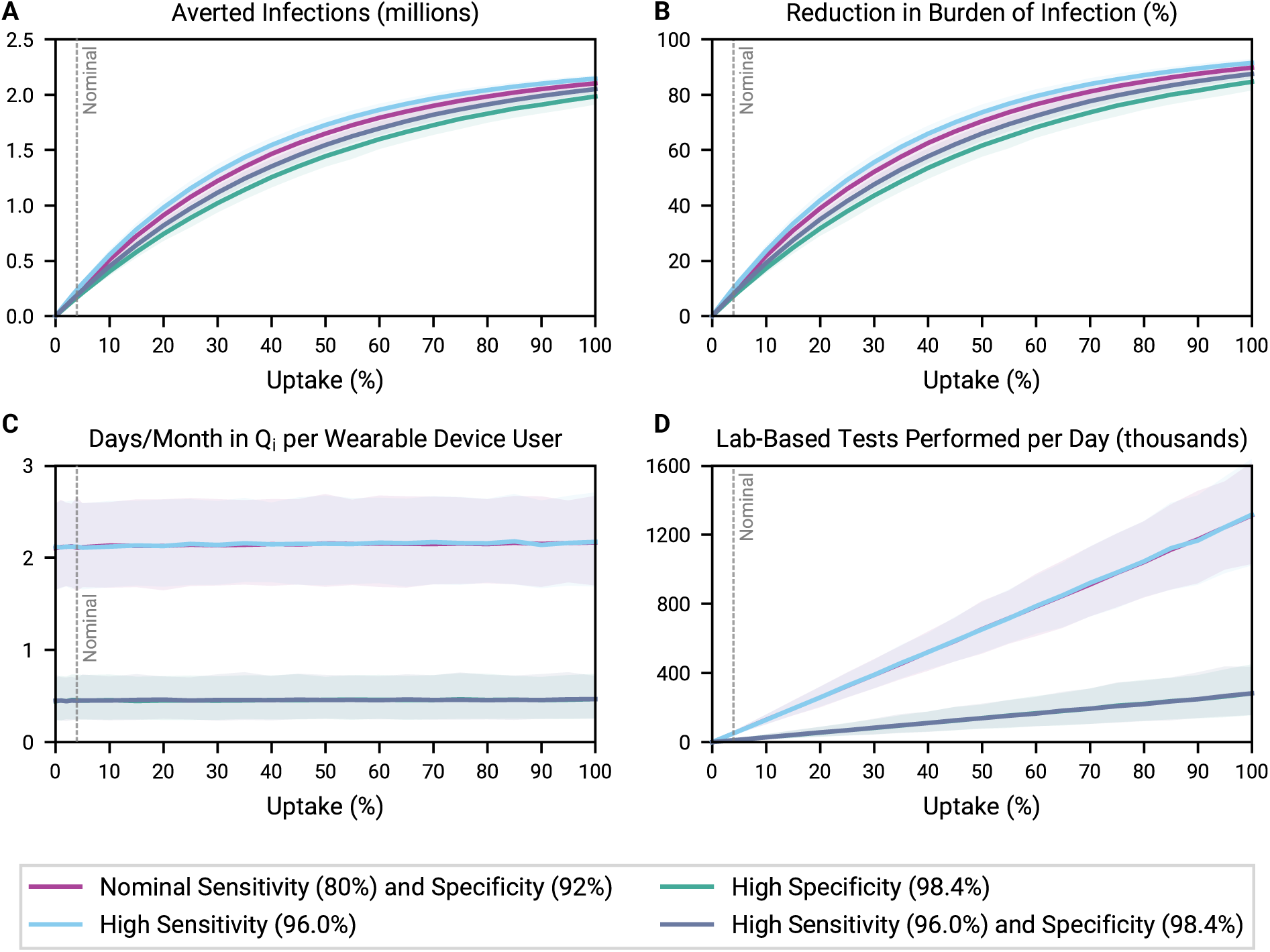
Impact of increasing uptake under different technology assumptions. Averted infections (A), reduction in the burden of infection (B), days incorrectly spent in quarantine per month per device user (C), and average daily demand for lab-based tests (D), all over the entire simulation period, as a function of increasing uptake. Grey dashed lines denote nominal uptake (4%). In the High Sensitivity and High Specificity scenarios, detection specificity and sensitivity are kept at their nominal values, respectively. In (C) and (D), the “Nominal Sensitivity and Specificity” and “High Sensitivity” curves overlap, and the “High Specificity” and “High Sensitivity and Specificity” curves overlap.

In all technology scenarios, increasing uptake averted more infections, though with diminishing returns (Figures 4a and 4b). As expected, improving detection specificity resulted in fewer averted infections when uptake is held constant; this effect was most pronounced at ∼30% to ∼60% uptake. The number of days incorrectly spent in quarantine per month per device user remained constant as a function of uptake but decreased from ∼2.15 to ∼0.45 when detection specificity was increased (Figure 4c). This ∼80% decrease was consistent with our definition of “high specificity” demonstrating that detection specificity directly influences the burden of incorrect quarantines on device users. The average daily demand for lab-based tests scaled linearly with uptake, but at a slower rate with improved detection specificity (Figure 4d).

### Impact of increasing adherence

Adherence to public health guidelines also impacts the success of pandemic control measures. Targeted policies – for example, compensating individuals in self-isolation – could help improve compliance with public health recommendations^20^. Here, we explored the role of adherence in wearable sensor deployment strategies (Figure 5; Supplementary Figure 1). We chose to present outcomes at all levels of adherence because compliance with public health guidelines could vary greatly, for example, from as low as 14% to as high as 86% (Supplementary Table 2). We kept uptake constant at 4% and assessed multiple technology scenarios using the same definitions of “high” detection sensitivity and specificity as above.

**Figure 5:**
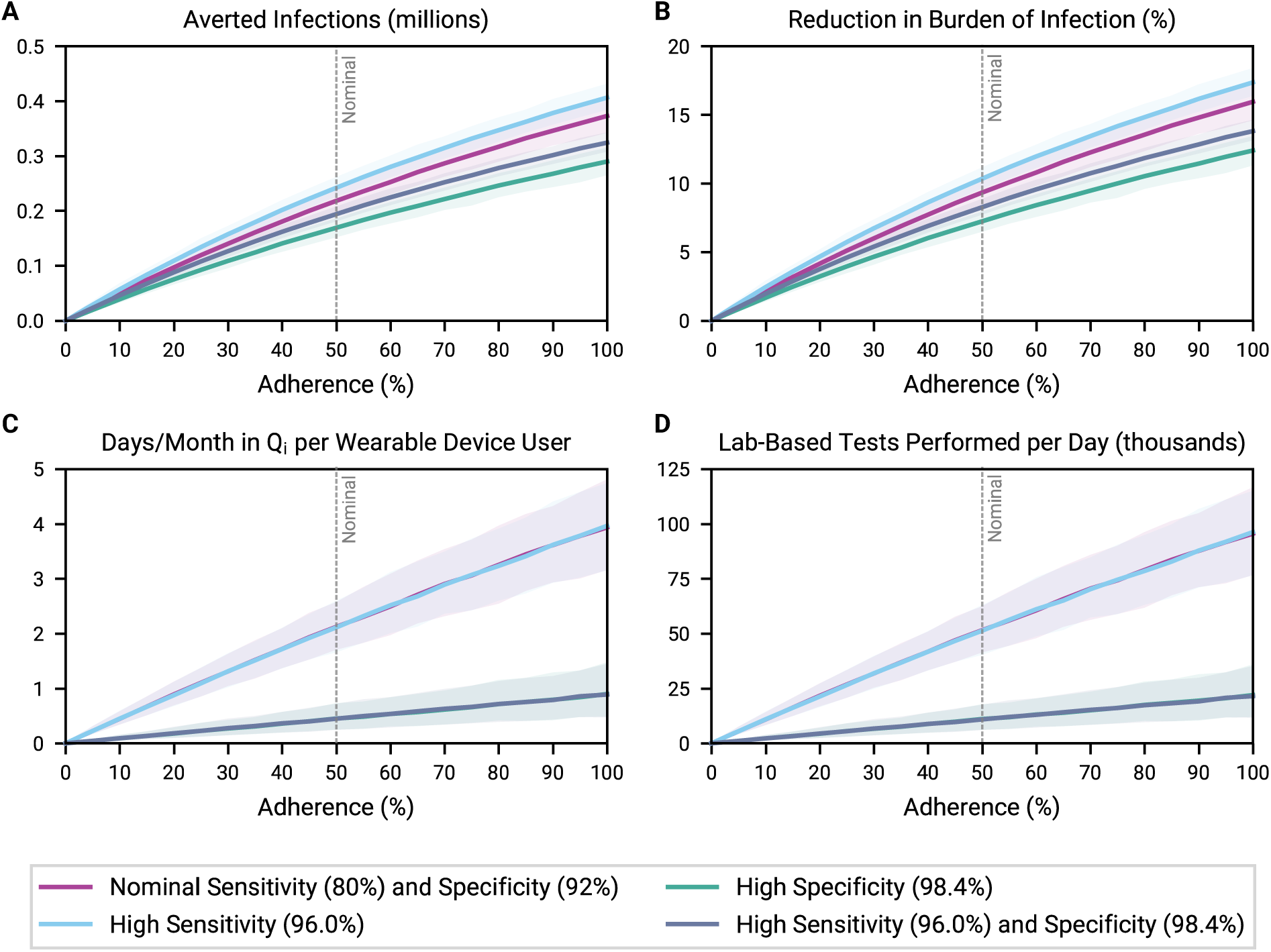
Impact of increasing adherence under different technology assumptions. Averted infections (A), reduction in the burden of infection (B), days incorrectly spent in quarantine per month per device user (C), and average daily demand for lab-based tests (D), all over the entire simulation period, as a function of increasing adherence. Grey dashed lines denote nominal uptake (4%). In the High Sensitivity and High Specificity scenarios, detection specificity and sensitivity are kept at their nominal values, respectively. In (C) and (D), the “Nominal Sensitivity and Specificity” and “High Sensitivity” curves overlap, and the “High Specificity” and “High Sensitivity and Specificity” curves overlap.

Adherence meaningfully impacted the burden of infection (Figure 5b). For example, with nominal detection sensitivity and specificity, increasing adherence among participating wearable device users from 20% to 80% tripled the achieved reduction in the burden of infection, raising it from 4.2% (95% CI: 3.6–4.7%) to 13.6% (95% CI: 12.4–14.8%). However, increasing the proportion of users who comply with notifications also magnified the consequences of false positive notifications: the number of days incorrectly spent self-isolating per month per user (Figure 5c) and the demand for lab-based tests (Figure 5d) grew proportionally with adherence. These social and resource costs grew at a slower rate with improved detection specificity.

### Impact of offering confirmatory rapid antigen tests

Our earlier findings suggested that false positive notifications of potential infection were the primary cause of unnecessary quarantines and lab-based tests. Improving detection specificity was one way to decrease false positive notifications. Here, we investigated whether offering confirmatory rapid antigen tests to users with a positive notification could also contribute to reducing unnecessary quarantines and lab-based tests (Figure 6; Table 2; Supplementary Table 6). We considered multiple scenarios, each with either low levels of uptake (0.5%) or adherence (14%), nominal levels of uptake (4%) or adherence (50%), or high levels of uptake (12.5%) or adherence (86%). We examined these scenarios in the cases of nominal detection sensitivity and specificity, and of “high” detection sensitivity and specificity (using the same definitions of “high” as above).

**Table 2:**
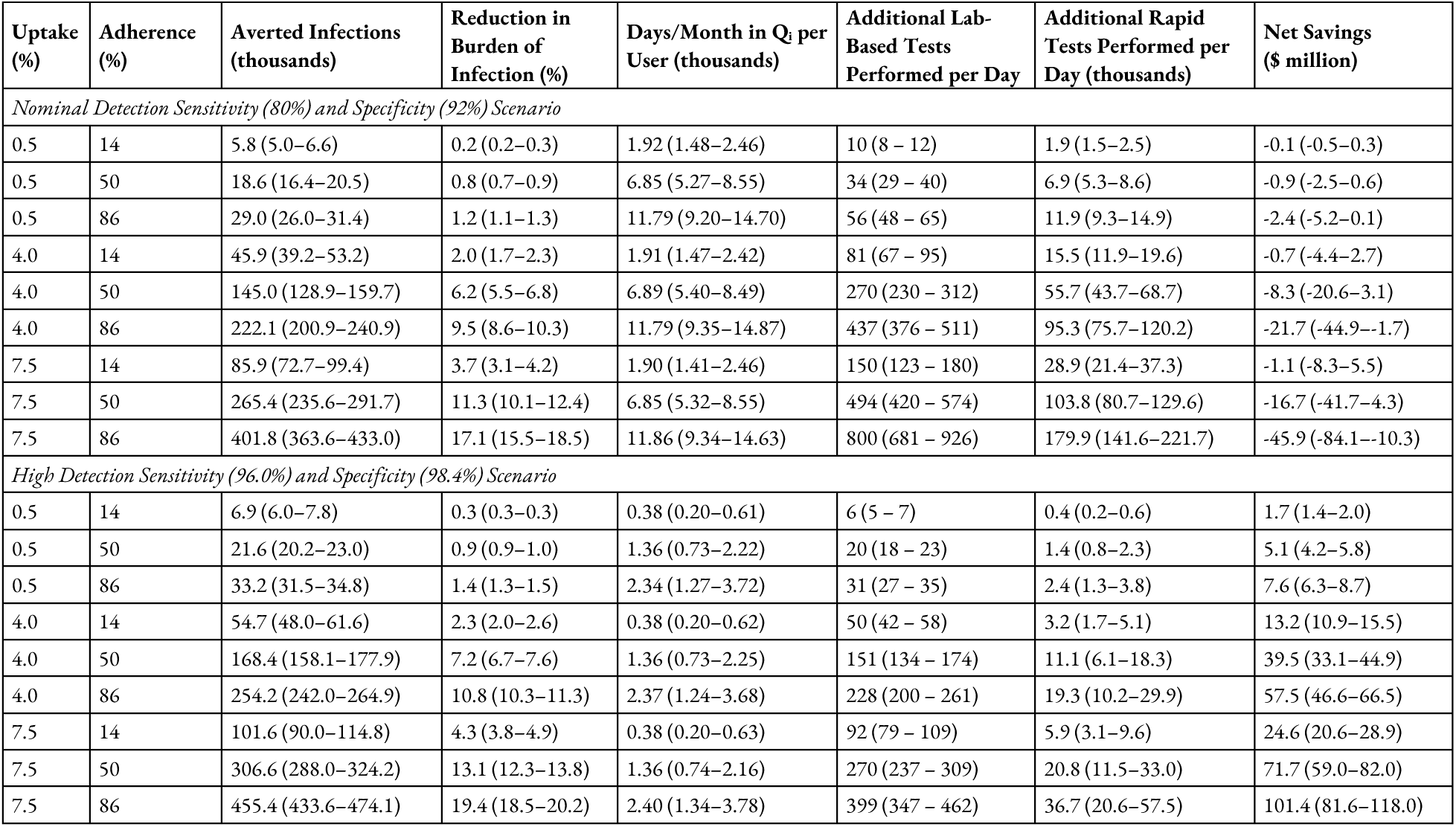
Impact of offering confirmatory antigen tests under different technology assumptions. 95% confidence intervals are listed in parentheses. Supplementary Table 6 depicts outcomes in analogous scenarios without rapid antigen tests.

**Figure 6:**
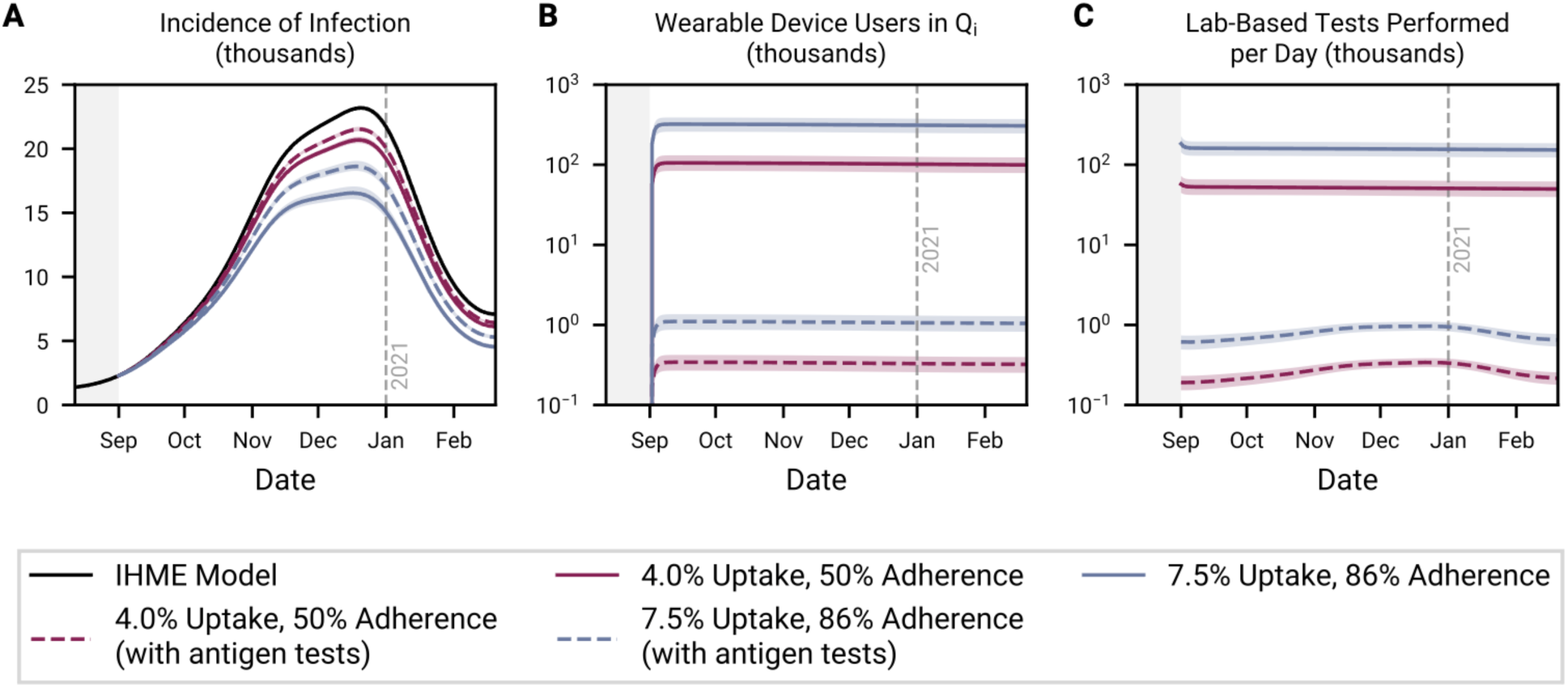
Wearable sensor deployment with confirmatory antigen tests. Time series depiction of the incidence of infection (A), the number of wearable device users incorrectly in quarantine (B), and the daily demand for lab-based tests (C). Detection sensitivity and specificity are set to their nominal values of 80% and 92%, respectively.

The use of antigen tests reduced the number of days incorrectly spent in quarantine by ∼300-fold by increasing the “effective specificity” of the strategy (Figure 6b). That is, with antigen tests, the likelihood of a *Susceptible* user being incorrectly prompted to quarantine on a given day fell from (1 – ν_w_) to the product of (1 – ν_w_) and (1 – ν_a_), where ν_w_ and ν_a_ are detection algorithm specificity and antigen test specificity, respectively. In earlier scenarios (Figures 4a and 5a), averted infections were decreased by improving detection specificity more than they were increased by improving detection sensitivity; fewer infections were averted in the high detection sensitivity and specificity scenarios than in the nominal detection sensitivity and specificity scenarios. Here, the specificity contributed by the antigen tests diminished the relative impact of improving detection specificity on averted infections: the “effective specificity” of the strategy was 99.976% with nominal detection specificity and 99.995% with high detection specificity (Supplementary Table 2)^21^. Instead, improving detection sensitivity was what increased the number of averted infections. Importantly, antigen tests had the secondary effect of decreasing the strategy’s “effective sensitivity” – the product of antigen test sensitivity (91.1%) and detection algorithm sensitivity^21^.

Offering confirmatory rapid antigen tests also decreased the demand for lab-based tests by ∼200-fold, providing the double benefit of alleviating the burden on testing infrastructure and decreasing costs (Figure 6c). We earlier found that in a baseline scenario (4% uptake, 50% adherence, 80% detection sensitivity, 92% detection specificity), between ∼40,000 and ∼65,000 additional lab-based tests would be required each day (Figure 2c). Here, in an analogous scenario, average daily demand dropped to 270 (95% CI: 230–312) additional lab-based tests, with 55,700 (95% CI: 43,700–68,700) antigen tests being performed each day instead (Table 2). Antigen tests were also more cost-effective tools than lab-based tests for confirming *Susceptible* users with false positive notifications were not infectious. Accounting for the costs of lab-based tests and savings associated with averted hospitalizations, we approximated that a baseline scenario would involve $825.7 (95% CI: $645.2– 1,035.7) million in net healthcare expenditures (Supplementary Table 6). Here, we approximated that the analogous scenario would involve $8.3 (95% CI: −3.1–20.6) million in net expenditures (Table 2). Unnecessary lab-based tests drove this ∼10-fold difference in costs.

## DISCUSSION

We used a counterfactual model of Canada’s second COVID-19 wave to demonstrate that wearable sensors capable of detecting infectious diseases before or absent symptoms can be useful tools for pandemic mitigation. Through continuous and non-invasive monitoring of physiological parameters, these devices can conceivably help FTTI systems identify hidden infection chains with minimal delay and without active user engagement or broad sharing of user data. We showed that (1) deploying today’s detection algorithms could have meaningfully reduced the second wave burden of infection, but with substantial social and resource costs; (2) improving detection algorithm specificity and offering confirmatory rapid antigen tests can help minimize unnecessary quarantines and lab-based tests; and (3) once false positive notifications are minimized, increasing uptake and adherence become effective strategies to scale the number of averted infections.

In theory, wearable sensor deployment reduces the burden of infection by decreasing the pool of infectious individuals (a function of detection algorithm sensitivity). Here, we found that detection specificity played an unexpectedly large role as well, with false positive notifications of potential infection prompting unnecessary quarantines and thereby decreasing the pool of susceptible individuals. Thus, although prioritizing uptake and adherence as part of a wearable sensor deployment strategy could mitigate a substantial number of infections, the unsustainable growth of associated costs should also be considered. In a baseline scenario, without improvements to detection specificity, every user would spend over two days a month on average incorrectly quarantining, and ∼40,000 to ∼65,000 additional confirmatory lab-based tests would be required each day. The social and economic harm caused by solely promoting uptake or adherence without improvements to detection specificity would likely undermine public confidence in and compliance with a wearable-based pandemic mitigation strategy^22^. Alavi *et al* found that many false positives were due to the detection algorithm identifying lifestyle-driven changes in resting heart rate (e.g., after intense exercise or alcohol consumption); accounting for these factors using more advanced algorithms may be one way to target improved detection specificity^10^.

We found that the inclusion of confirmatory antigen testing was a valuable mechanism, beyond improving detection specificity, to increase the “effective specificity” of the strategy and decrease the overall false positive rate. The inclusion of antigen testing decreased days incorrectly spent in quarantine by ∼300-fold and brought the additional demand on lab-based testing infrastructure to justifiable levels. In general, antigen tests were more cost-effective and immediate tools than lab-based tests to confirm a *Susceptible* device user was not infectious. However, even with the inclusion of antigen tests, improvements to detection specificity still had value. Relative to scenarios with nominal detection specificity, with “high” detection specificity, we observed a ∼4-fold reduction in days incorrectly spent in quarantine per month per user, a ∼2-fold reduction in lab-based tests performed each day, and a ∼5-fold reduction in antigen tests used each day. Importantly, a strategy in which antigen tests support the deployment of wearable sensors is notably different from one involving frequent use of rapid antigen tests for surveillance testing^23^. On their own, broad antigen test-based screening approaches require tremendous manufacturing volumes, infrastructure, and funding^24^. Conversely, wearable sensors can non-invasively detect infections without active user engagement, reducing the effort required to participate. Further, in the scenarios explored here, detection algorithm specificity governed how efficiently rapid antigen tests were deployed.

Our work has important limitations. First, we assumed that SARS-CoV-2 epidemiology and wearable device use were homogenous within the population. Our determination of the transmission rate from epidemiological data inherently results in a “well-mixed” approximation for this value averaged over population-level heterogeneities such as age and super-spreading. As well, the COVID-19 pandemic has disproportionately impacted low-income and minority groups, while younger and wealthier individuals are more likely to own wearable devices^25,26^. Future studies could consider a policy where the incentive to download the application is increased to become a subsidy for purchasing a wearable device, reducing the participation barrier. Second, we used an existing model for the incidence of infection which has its own assumptions and limitations^13^. Third, we made the simplifying assumption that all users without symptoms (and that no users with symptoms) could benefit from wearable-informed prompts to take a confirmatory test and self-isolate. Fourth, we did not consider how uptake or adherence might vary over time or with detection accuracy^18,22^. Fifth, we used median values for SARS-CoV-2 infection parameters (e.g., latent period) and did not account for reinfections or vaccinations. Finally, we did not account for administrative costs or second order savings (e.g., avoided worker’s compensation payouts for 14-day quarantines).

To our knowledge, this is the first study to explore the real-world impact of deploying wearable sensors for pandemic mitigation. Using the example of COVID-19, we demonstrated that these devices have the potential to support FTTI systems with real-time detection of presymptomatic and asymptomatic infections, and ultimately reduce the burden of infection. However, in cases with insufficient detection algorithm specificity, complementary interventions that reduce false positives are required to minimize social costs and resource demands. In the future, there is clear merit to further exploring how wearable sensors can be incorporated into FTTI systems for pandemic mitigation, and whether these devices continue to have public health utility once an endemic phase is reached, complementary to vaccines.

## Data Availability

All data produced in the present work are contained in the manuscript.

## Acknowledgements

We would like to thank Chadi M. Saad-Roy (Princeton University) and Étienne Racine (McGill University) for guidance with modeling, Andrew Soltan (Oxford University) for insightful discussions, and Arash Alavi (Stanford University) for assistance with data interpretation.

## STATEMENTS

### Author Contributions

ND and GB contributed to the conceptualization; ND, RKA, GB, MW, and CEW contributed to the methodology; ND and GB contributed to the writing the original draft; ND contributed to software; ND and RKA contributed to visualization; ND, RKA, GB, MW, MPS, JRC, and CEW contributed to review and editing; ND, JRC, and CEW contributed to supervision.

### Materials and Correspondence

Correspondence and material requests should be addressed to Nathan Duarte and Caroline E. Wagner.

## Supplementary Appendix

### 1. Modeling assumptions, variables, and equations

**Supplementary Table 1:** SARS-CoV-2 characteristics. We obtained the presymptomatic infectious period by subtracting the latent period (3.69 days) from the incubation period (5 days)^1,2^. We obtained the asymptomatic and symptomatic infectious period by subtracting the presymptomatic infectious period (1.31 days) from the total infectious period (7 days)^1–3^. During Monte Carlo simulations, we modeled asymptomatic prevalence as a beta random variable with a mean of 0.4 and a sample size of 200; a sample size of 200 is greater than 75% of the study populations examined by Oran and colleagues^4^.

| Parameter | Symbol | Value |
| --- | --- | --- |
| Latent period (exposure to infectiousness) | $1 / \alpha$ | 3.69 days<sup>1</sup> |
| Presymptomatic infectious period | $1 / \tau$ | 1.31 days<sup>1,2</sup> |
| Asymptomatic and symptomatic infectious period | $1 / \gamma$ | 5.69 days<sup>1-3</sup> |
| Asymptomatic prevalence | $\rho$ | 40%<sup>4</sup> |
| Transmission potential without symptoms relative to with symptoms | $\lambda$ | 55%<sup>1</sup> |
| Daily new infections | $\pi$ | varies |
| Average number of transmissions per infectious person per day | $\beta$ | varies |

**Supplementary Table 2:** Wearable device and policy parameters.

| Parameter | Symbol | Nominal Value | Notes |
| --- | --- | --- | --- |
| Wearable device uptake | $\theta$ | 4% | We defined uptake as the percent of the population that owns a wearable device, has downloaded the detection/notification application, *and* uses the device enough to collect sufficient data for detection. We estimated uptake would range from 0.5% to 7.5% at baseline. Supplementary Tables 3 and 4 provide an example of this calculation.<br><u>Device ownership</u>: Estimates from 2018 place device ownership in Canada between 22% and 25%<sup>5,6</sup>.<br><u>Download rate</u>: A study found that the baseline download rate of Germany's national contact tracing application was between ~8% and ~11%; incentives could increase this rate<sup>7</sup>. Paré and colleagues found that 57% of owners regularly track their health with their device<sup>5</sup>. Here, we looked at download rates ranging from 10% to 60%.<br><u>Utilization</u>: In research studies where wearable devices were used to track health, usage rates ranged from as low as 24%, to 50% in the long run<sup>8,9</sup>. |
| Adherence to wearable device notification | $\psi$ | 50% | We defined adherence as the proportion of wearable device users that comply with recommended next steps upon notification of potential infection. At baseline, next steps include seeking a confirmatory lab-based test, self-isolating while awaiting a result, and quarantining until recovery if the result is positive. With antigen tests, compliant users also take a confirmatory rapid antigen test prior to seeking a lab-based test. In Israel, at least ~53% of antigen test kit results were reported (~613,000 reports out of ~1,150,000 kits taken home)<sup>10,11</sup>. In Norway, up to 70% of those with a suspected diagnosis and up to 86% with a positive diagnostic test adhered to quarantine<sup>12</sup>. However, adherence could be as low as ~14%, if one considers Canada's COVID Alert contact tracing app's reporting rate in light of confirmed cases as of July 27, 2021<sup>13</sup>. During Monte Carlo simulations, we modeled adherence as a beta random variable with a mean of 0.5 and a sample size of 1723; 1723 was the sample size for the relevant experiment in the Norway study. |
| Detection algorithm sensitivity | $\sigma_w$ | 80% | We defined sensitivity as the proportion of infected yet asymptomatic wearable device users (i.e., *Exposed*, *Presymptomatic*, and *Asymptomatic*) who receive a notification of potential infection prior to recovering and entering the *Removed* compartment. Alavi and colleagues' NightSignal algorithm achieved a sensitivity of ~80%, which is a plausible value based on other efforts to develop similar algorithms<sup>14,15</sup>. During Monte Carlo simulations, we modeled sensitivity as a beta random variable with a mean of 0.80 and a sample size of 84; 84 was the size of the sample used to calculate the NightSignal algorithm's sensitivity. |
| Detection algorithm specificity | $v_w$ | 92% | We defined specificity as the probability that, on a given day, a healthy (i.e., *Susceptible*) user does not receive a notification of potential infection. Alavi and colleagues' NightSignal algorithm gave potentially healthy users 0.0819 false positive notifications ("red alerts") per day on average, corresponding to a specificity of ~92%<sup>14</sup>. During Monte Carlo simulations, we modeled specificity as a beta random variable with a mean of 0.92 and a sample size of 818; alarm data from 818 potentially healthy users in Alavi and colleagues' dataset were used to calculate the false positive rate. |
| Detection algorithm sensitivity adjustment factor | $\kappa$ | $(1/\alpha + 1/\tau + 1/\lambda)^{-1}$ days<sup>-1</sup> | We assumed that $\sigma_w$ is applied uniformly across *Exposed*, *Presymptomatic*, and *Asymptomatic* states such that by the time infectiousness ends, $\sigma_w$ of infected users without symptoms are notified. |
| Detection algorithm specificity adjustment factor | $\chi$ | $(1)^{-1}$ days<sup>-1</sup> | We assumed that $\nu_w$ is applied over a period of one day such that on any given day, *Susceptible* users receive an incorrect (i.e., false positive) notification of potential infection with probability $(1-\nu_w)$. |
| Rapid antigen test sensitivity | $\sigma_a$ | 91.1% | We used the lowest performance reported for Abbot's Panbio test<sup>16</sup>. Independent evaluations of this test suggest lower sensitivity in presymptomatic and asymptomatic individuals<sup>17,18</sup>. However, we reasoned that the receipt of a wearable-informed notification of potential infection raised the pretest probability of infection, thereby improving the negative predictive value (NPV) of the tests<sup>19</sup>. We performed a sensitivity analysis on the sensitivity of rapid antigen tests (Supplementary Figure 5). |
| Rapid antigen test specificity | $\nu_a$ | 99.7% | We used the lowest performance reported for Abbot's Panbio test<sup>16</sup>. Independent evaluations suggest that this test's specificity is even higher<sup>17,18</sup>. |
| Lab-based test turnaround time | $\epsilon$ | 2 days | In general, 1-3 days based on Health Canada reporting<sup>20</sup>. |
| Transmission rate increase factor | $a$ | 1 | Used to test the scenario in which device users who do not receive a notification of potential infection act in a riskier fashion, effectively increasing their transmission rate<sup>21</sup>. |

Model equations, in addition to Equation (1), used to extract transmission rate (β) from the IHME infection model:

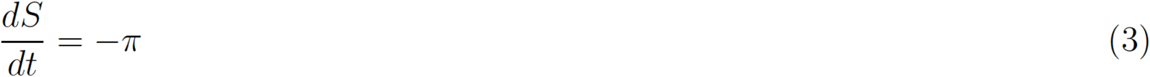

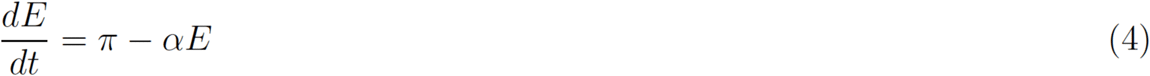

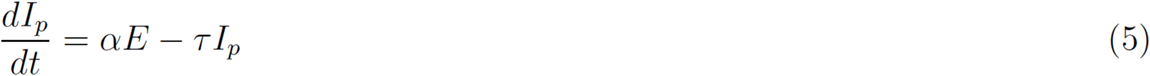

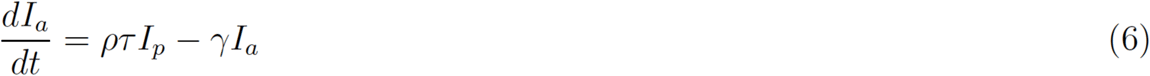

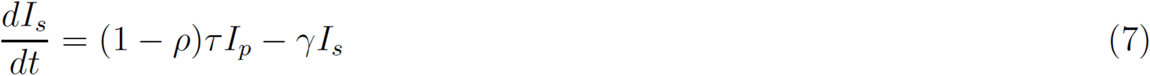

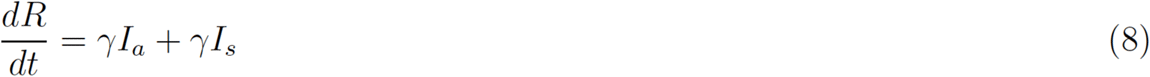

Model equations, in addition to Equation (2), used to run simulations:

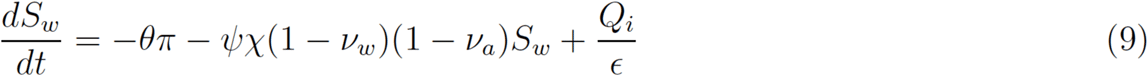

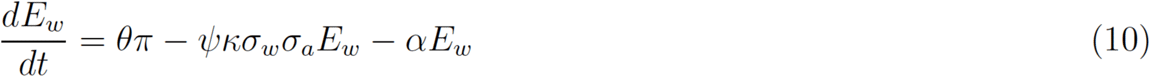

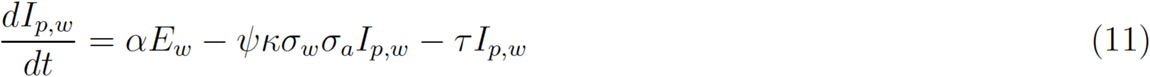

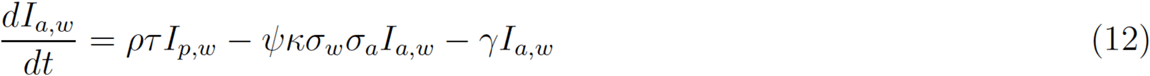

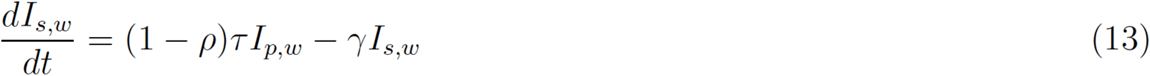

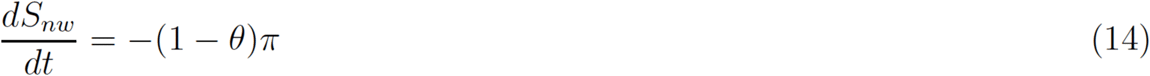

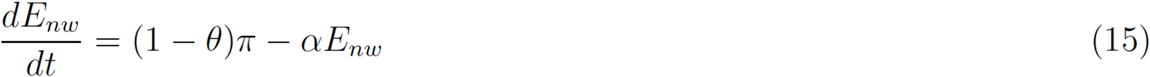

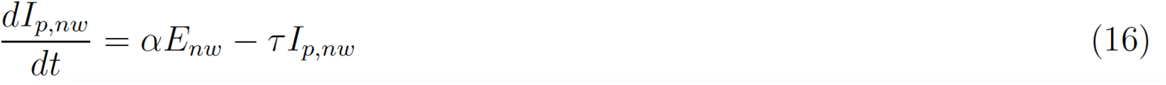

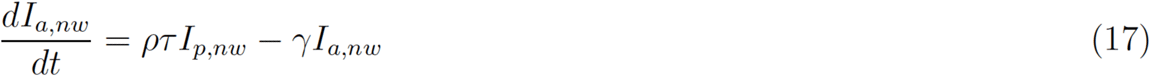

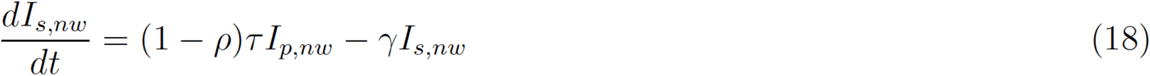

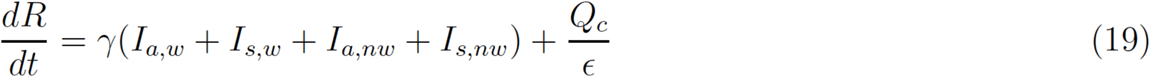

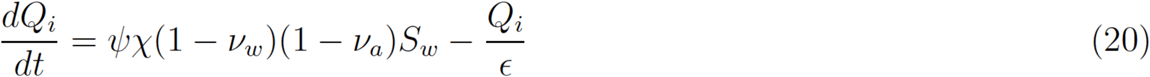

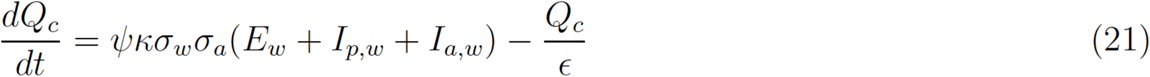

### 2. Estimating ranges for uptake

Below, Supplementary Tables 3 and 4 delineate how we estimated plausible ranges for uptake in a baseline scenario. First, we multiplied the download rate by the proportion of the population that owns a device to obtain a plausible range for the proportion of the population that owns the application (Supplementary Table 3). Then, we calculated a range for uptake by multiplying the proportion of individuals that own the application by expected levels of utilization (Supplementary Table 4). Accounting for utilization was a necessary step because not all individuals who download the application to their device use it enough to provide sufficient data for the algorithm to function correctly^14^. We concluded that in a baseline scenario, uptake would likely range from 0.5% to 7.5%. Assumptions around parameter values are listed in Supplementary Table 2 above.

**Supplementary Table 3:**
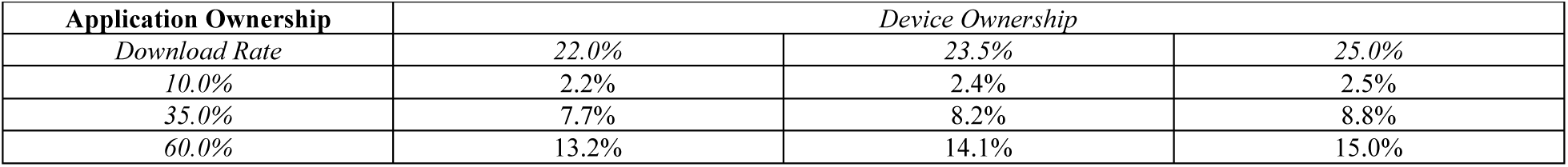
Calculation of the proportion of individuals who own the application.

| **Application Ownership** | *Device Ownership* |  |  |
| --- | --- | --- | --- |
| *Download Rate* | *22.0%* | *23.5%* | *25.0%* |
| *10.0%* | *2.2%* | *2.4%* | *2.5%* |
| *35.0%* | *7.7%* | *8.2%* | *8.8%* |
| *60.0%* | *13.2%* | *14.1%* | *15.0%* |

**Supplementary Table 4:** Calculation of the proportion of individuals who own and use the application.

| **Uptake** | *Utilization* |  |  |
| --- | --- | --- | --- |
| *Application Ownership* | *24.0%* | *37.0%* | *50.0%* |
| *2.2%* | *0.5%* | *0.8%* | *1.1%* |
| *8.6%* | *2.1%* | *3.2%* | *4.3%* |
| *15.0%* | *3.6%* | *5.6%* | *7.5%* |

### 3. Estimating economic impact

To generate a first-order estimate of net healthcare expenditures in particular scenarios, we subtracted savings from expenditures. Savings included costs avoided from averting hospitalizations. Expenditures included the costs of lab-based tests and antigen tests, where applicable. We calculated the number of averted hospitalizations and tests from our simulations using Equations (22) to (24) below. We multiplied these volumes by the costs in Supplementary Table 5, as in Equation (25), to generate a first-order approximation of net healthcare expenditures.

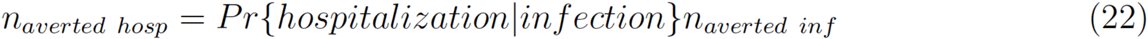

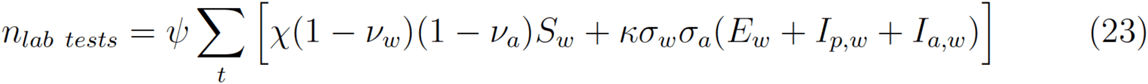

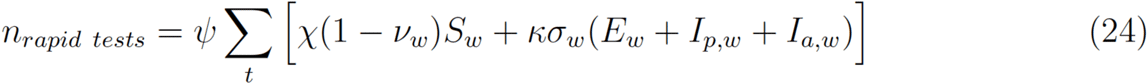

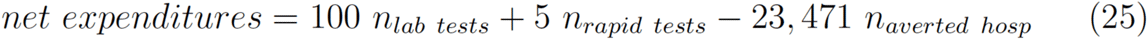

**Supplementary Table 5:** Costs used in economic impact calculations and associated assumptions.

| Parameter | Value | Notes |
| --- | --- | --- |
| Average cost of a COVID-19 hospitalization | \$23,471 | Average cost between January 2020 and March 2021; includes normal visits and ICU visits<sup>22</sup>. Data downloaded on December 16, 2021. |
| Infection hospitalization ratio | 1.31% | We calculated the infection hospitalization ratio by dividing the total number of COVID-19 hospitalizations by the total number of infections, all within the simulation period<sup>23</sup>. We offset the hospitalization counts by 7 days to align them with infections. |
| Cost of an antigen test | \$5 | Based on rates achieved when tests are purchased at scale<sup>24</sup>. |
| Cost of a diagnostic test | \$100 | Large variation in this figure; used rule of thumb<sup>25</sup>. |

### 4. Additional results

**Supplementary Figure 1:**
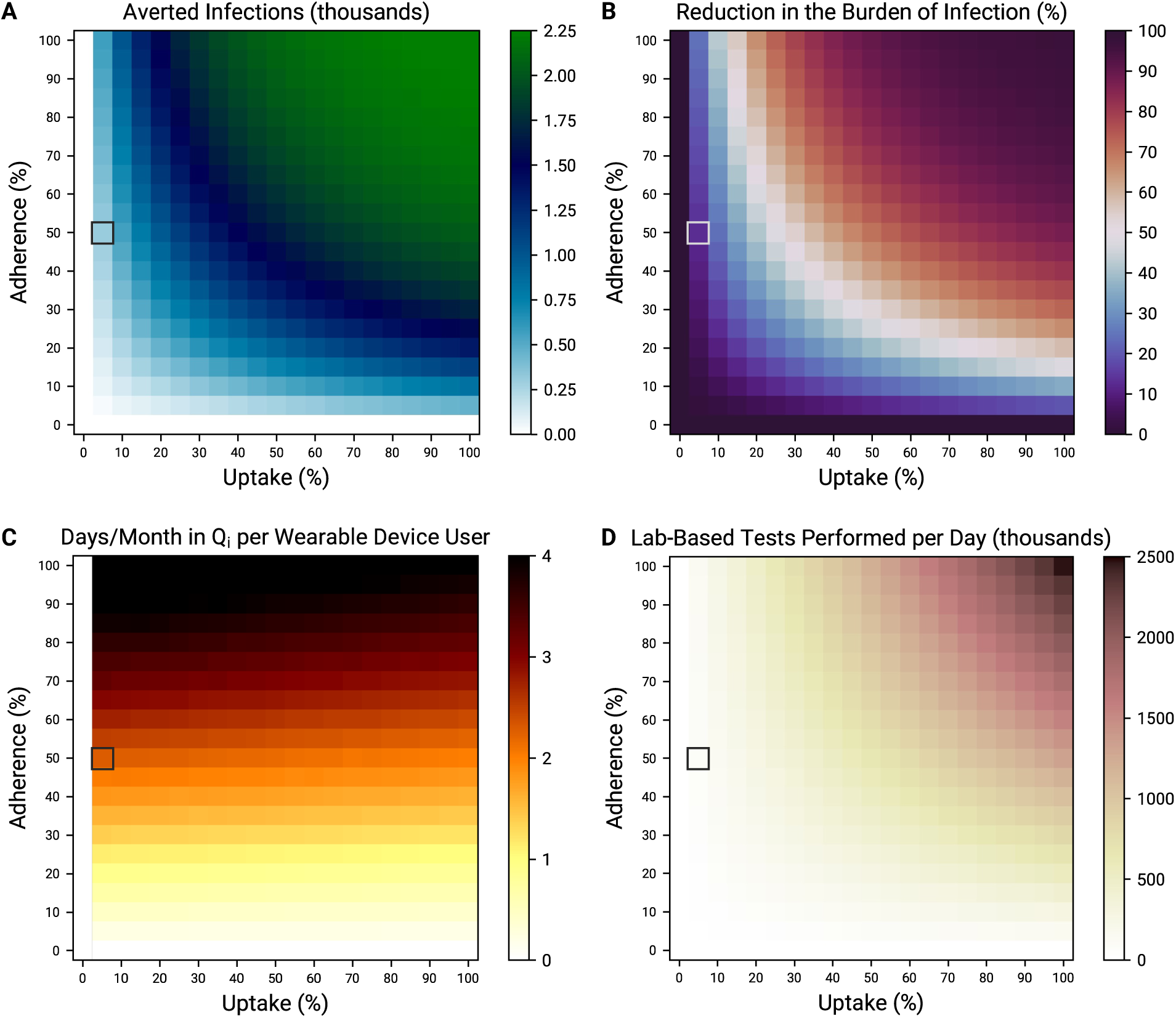
Impact of simultaneously increasing uptake and adherence. Averted infections (A), reduction in the burden of infection (B), days incorrectly spent in quarantine per month per device user (C), and average daily demand for lab-based tests (D), all over the entire simulation period, as a function of uptake and adherence. Grey boxes denote nominal sensitivity (80%) and specificity (92%).

**Supplementary Table 6:**
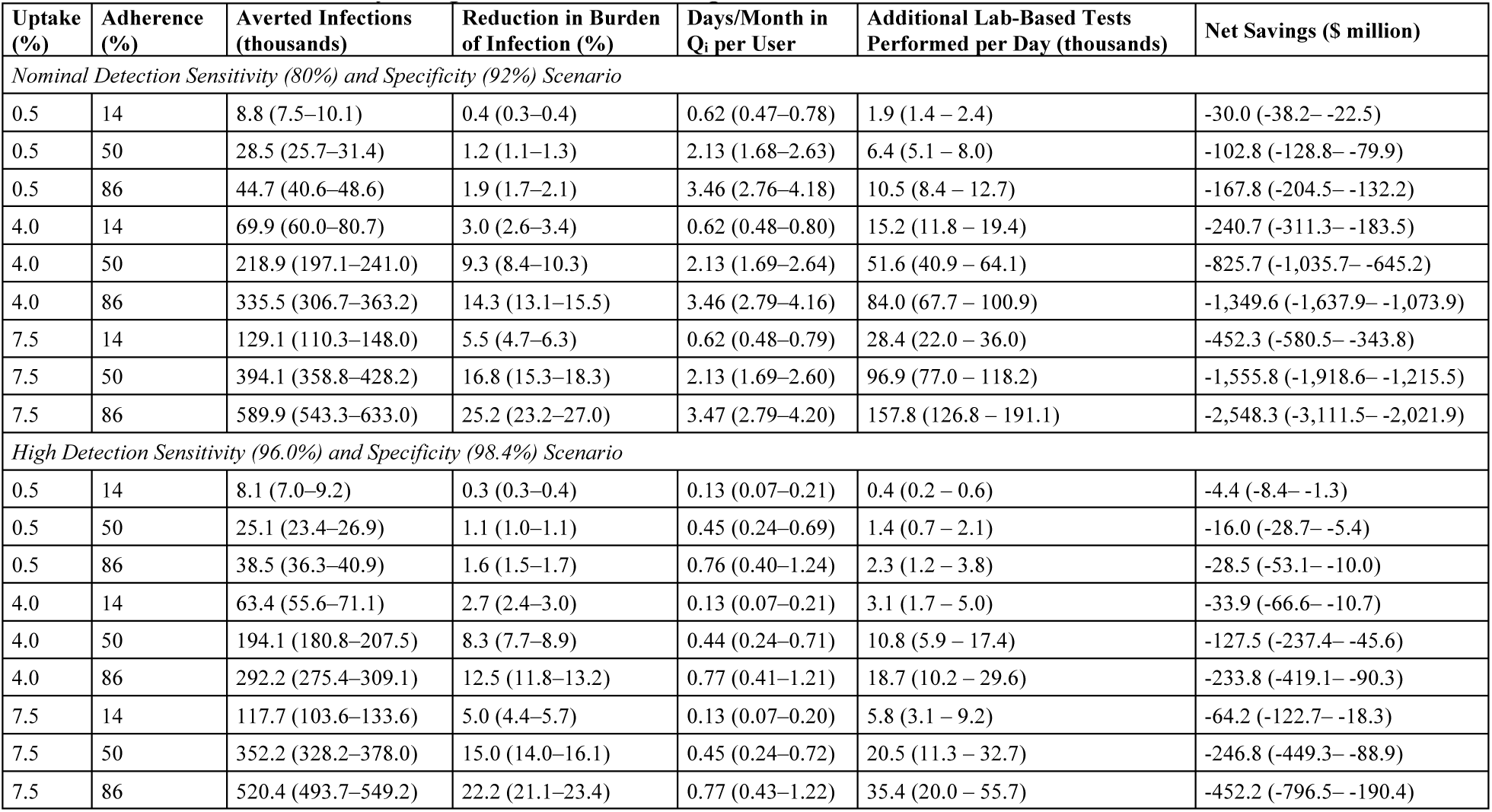
Wearable sensor deployment scenarios under different technology assumptions. 95% confidence intervals are listed in parentheses. This table serves as a counterpart to Table 2 so that analogous scenarios with and without confirmatory antigen tests can be compared.

### 5. Sensitivity analyses

First, we looked at whether, instead of remaining constant, the transmission rate of device users who do not receive a notification of potential infection might in fact increase due to a sense of false confidence (Supplementary Figure 2)^21^. We modulated *a* in Equation (2) to do so; a 5% increase in the transmission rate, for example, meant *a* was set to 1.05. Increase in transmission among device users relative to historical levels reduced the number of averted infections. The number of incorrect quarantines was not impacted. The implication of this finding is that public health leaders would need to communicate the limitations of wearable sensors with respect to detecting infections and emphasize that a lack of a notification does not rule out potential infection.

**Supplementary Figure 2:**
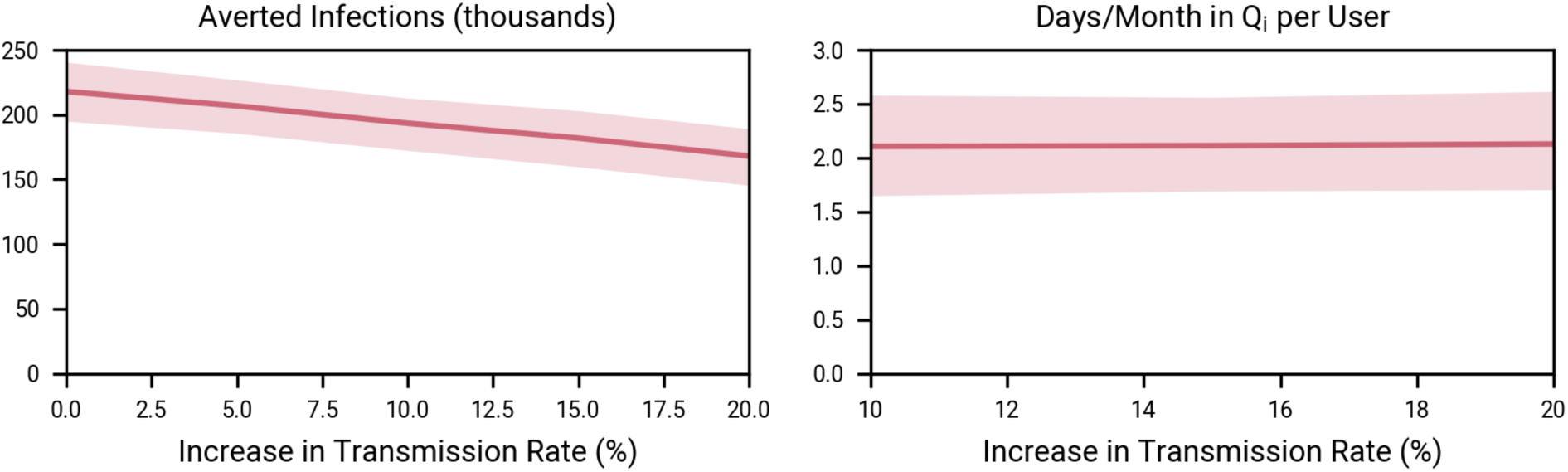
Impact of increased transmission among device users who are not notified of potential infection in a baseline scenario. We assumed 4% uptake, 50% adherence, and that transmission among non-users was unchanged.

Second, we looked at the impact of asymptomatic prevalence (Supplementary Figure 3). It was important to perform this analysis because our model only accounts for notifications sent to presymptomatic and asymptomatic individuals, yet there is still a lack of consensus on a specific value for the asymptomatic prevalence^4^. As expected, with greater asymptomatic prevalence, more individuals could benefit from wearable device use and more infections could be averted. The success of this strategy was not dependent on a particular value of asymptomatic prevalence. The number of incorrect quarantines was not impacted.

**Supplementary Figure 3:**
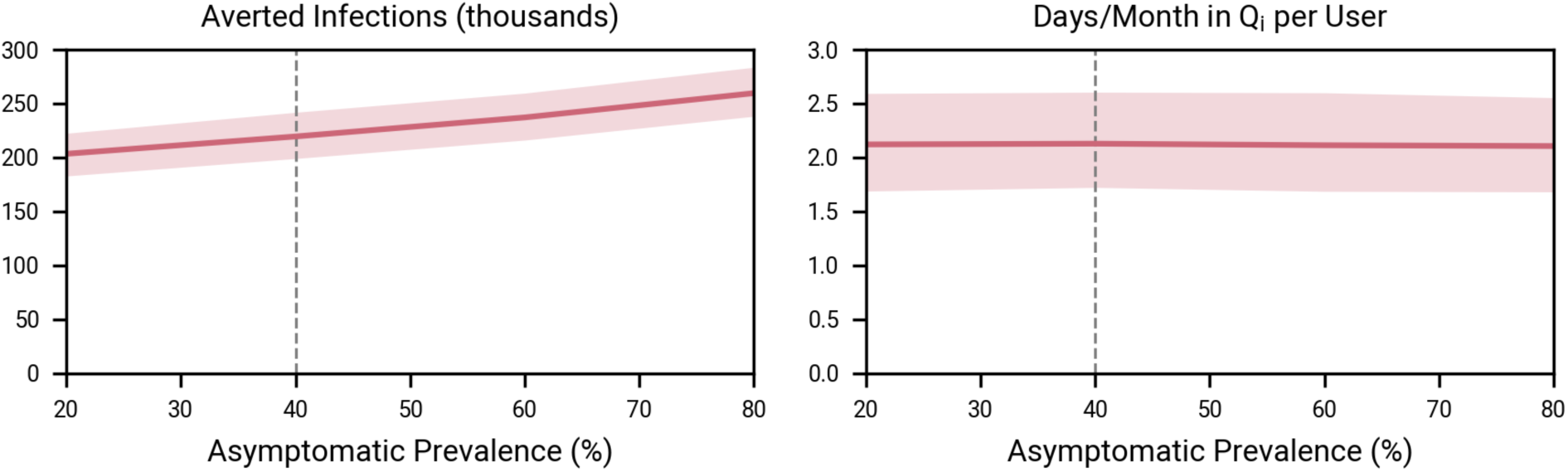
Impact of asymptomatic prevalence in a baseline scenario. We assumed 4% uptake and 50% adherence. We continued to model asymptomatic prevalence as a beta-distributed random variable (Supplementary Table 1). The dashed grey line represents nominal asymptomatic prevalence (40%).

Third, we investigated how the decrease in the number of *Susceptible* individuals resulting from incorrect quarantines influenced the number of averted infections in a baseline scenario (Supplementary Figure 4). We compared the number of averted infections with nominal (92%) and perfect (100%) detection specificity as detection sensitivity increased. In a baseline scenario, a meaningful proportion – 28.6% (95% CI: 18.3–38.2%) in the case of nominal detection sensitivity – of averted infections were driven by incorrect quarantines. This proportion decreased with increasing sensitivity. Consistent with earlier findings, it remains important to minimize unnecessary quarantines – social costs can be substantially reduced while still averting a meaningful number of infections, especially if detection sensitivity can be improved in parallel.

**Supplementary Figure 4:**
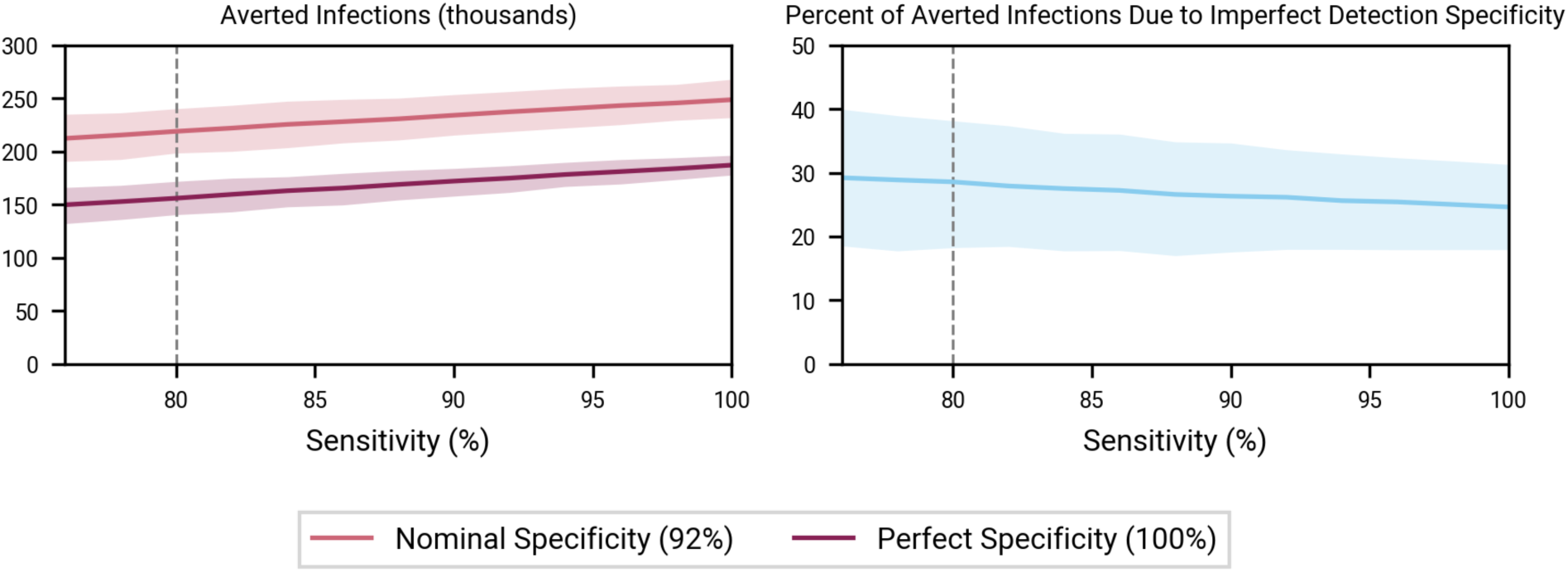
Impact of incorrect quarantines on averted infections in a baseline scenario. We assumed 4% uptake and 50% adherence. The dashed grey line depicts nominal detection algorithm sensitivity (80%).

Fourth, we investigated the impact of lower rapid antigen test sensitivity (Supplementary Figure 5). First, fewer infections were averted overall – as discussed, using antigen tests to minimize incorrect quarantines increased the pool of susceptible individuals. Second, averted infections grew linearly with test sensitivity: a ∼10% increase in test sensitivity resulted in a ∼14,000 increase in averted infections. These two effects result in a tradeoff between missing more infectious individuals (imperfect antigen test sensitivity) and decreasing false positive prompts to seek a lab-based test and self-isolate while awaiting the results (near perfect antigen test specificity). We believe the use of antigen tests as a complementary mechanism could be justified. Although fewer infections are averted relative to wearable sensor deployment without antigen tests, hundreds of thousands of infections are still averted relative to the counterfactual scenario – and in a resource-efficient and socially acceptable fashion. Certainly, infected individuals with a false negative antigen test might act in a riskier fashion – even still, there remains an opportunity to avert a substantial number of infections (Supplementary Figure 2). We also point out that improving detecting algorithm sensitivity could help counteract the effect of imperfect antigen test sensitivity (Table 2).

**Supplementary Figure 5:**
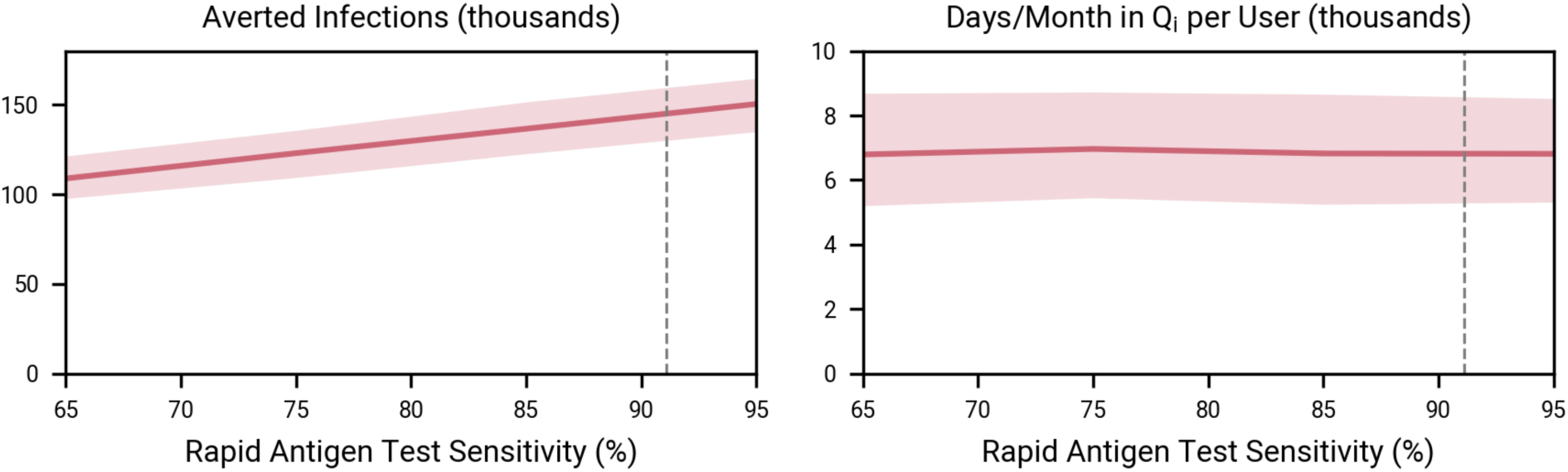
Impact of antigen test sensitivity. We assumed 4% uptake and 50% adherence. The dashed grey line represents nominal antigen test sensitivity (91.7%).

Fifth, we explored the impact of lab-based test turnaround time to determine whether minimizing this variable should be a policy priority (Supplementary Figure 6). Throughout this study, we set turnaround time to its nominal value of two days. With longer turnaround times, individuals incorrectly in quarantine would remain there longer, further decreasing the pool of *Susceptible* individuals. As expected, more infections are consequently averted. However, there are other mechanisms (e.g., improving detection sensitivity) for increasing averted infections that are not accompanied by as large of an increase to social costs. Thus, as is already the case outside the context of wearable sensor deployment, it would make sense to minimize turnaround time.

**Supplementary Figure 6:**
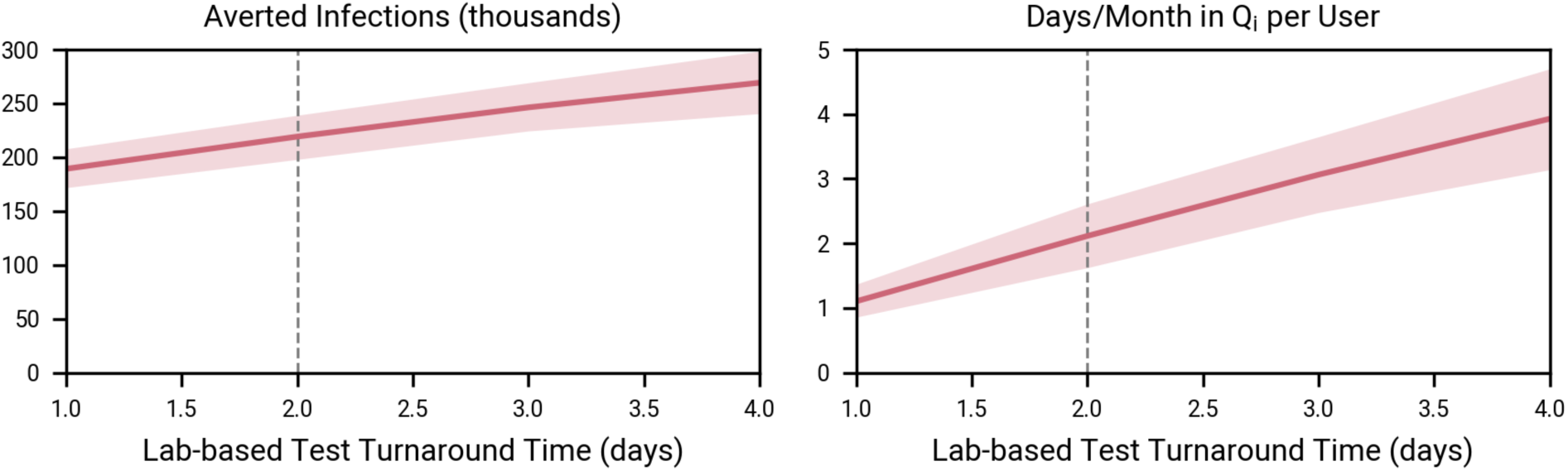
Impact of lab-based test turnaround time. We assumed 4% uptake and 50% adherence.

Finally, to understand the impact of detection algorithm sensitivity on the incidence of infection, we investigated the scenario in which device users are randomly asked to seek a lab-based test and self-isolate while awaiting the result (Supplementary Figure 7). We set the likelihood of being asked to quarantine to the false positive rate, still treating detection specificity as a beta-distributed random variable and setting it to its nominal value. As expected, the reduction in detection sensitivity substantially reduces the number of averted infections. The number of individuals incorrectly in quarantine is unimpacted because it is governed by detection specificity, not sensitivity.

**Supplementary Figure 7:**
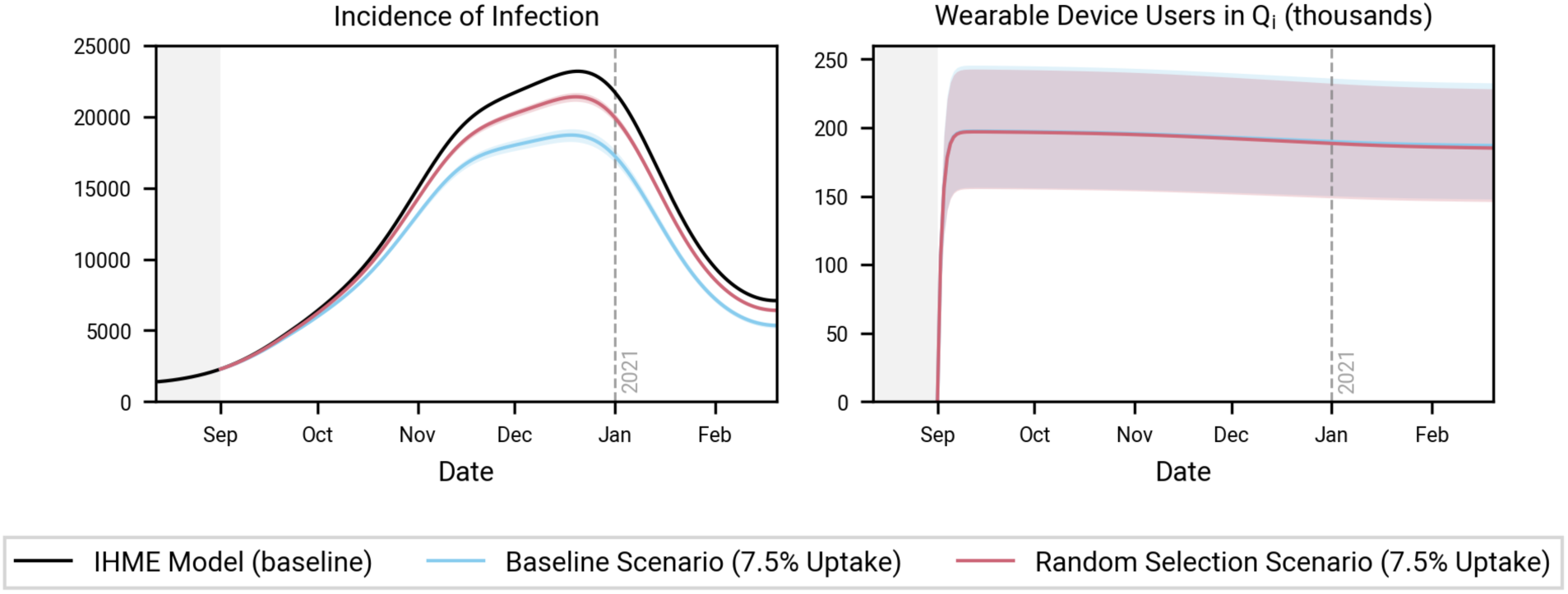
Impact of randomly asking device users to seek a lab-based test and quarantine while waiting for the result. We assumed 7.5% uptake, still with 50% adherence, to illustrate the relative difference in the incidence of infection.

